# Real-World Type 2 Diabetes Second-Line Treatment Allocation Among Patients

**DOI:** 10.1101/2025.03.26.25324631

**Authors:** Jaysón Davidson, Rohit Vashisht, Kendra Radtke, Ayan Patel, Suneil K. Koliwad, Atul J. Butte

**Affiliations:** Bakar Computational Health Sciences Institute, University of California San Francisco, San Francisco, CA; Center for Data-driven Insights and Innovation, University of California Health, Oakland, CA; Diabetes Center, University of California San Francisco, San Francisco, CA; Department of Medicine, Division of Endocrinology & Metabolism, University of California San Francisco, San Francisco, CA

**Author notes:** Corresponding Author: Atul J. Butte M.D., Ph.D.

**Keywords:** Type 2 diabetes, treatment decision-making, social determinants of health, healthcare disparities, area deprivation index

## Abstract

**Objective:** This study aimed to evaluate the impact of socioeconomic disparities on the allocation of second-line treatments among patients with type 2 diabetes (T2D).

**Materials and Methods:** We conducted an observational study using real-world data from over 9 million patients across five University of California Health centers. The study included patients who initiated a second-line T2D medication after metformin, with hemoglobin A1c (HbA1c) measurements within ±7 days of treatment initiation from 2012 through September 2024. Multinomial regression models assessed the association between socioeconomic status and second-line treatment choices. Additionally, we used the GPT-4 large language model with a zero-shot learning approach to analyze 270 clinical notes from 105 UCSF patients. GPT-4 identified adverse social determinants of health (SDOH) across six domains: transportation, housing, relationships, patients with children, support, and employment.

**Results:** Among 15,090 patients (56.7% male, 43.3% female; mean age 59.3 years; mean HbA1c 8.91%), second-line treatments included sulfonylureas (SUs; n = 6,732), DPP4 inhibitors (n = 2,918), GLP-1 receptor agonists (n = 2,736), and SGLT2 inhibitors (n = 2,704). Patients from lower socioeconomic neighborhoods were more likely to receive SUs over other medications: DPP4i (OR = 0.96, [95% CI, 0.95-0.98]), GLP-1RA (OR = 0.94, [95% CI, 0.92-0.96]), SGLT2i (OR = 0.95, [95% CI, 0.93-0.97]). In UCSF clinical notes, we identified adverse SDOH including housing (n=8), transportation (n=1), relationships (n=22), employment (n=12), support (n=1), and patients with children (n=25).

**Conclusions:** Socioeconomic factors influence second-line T2D treatment choices. Addressing these disparities is essential to ensuring equitable access to advanced T2D therapies.

## Introduction

The annual prevalence of Type 2 diabetes (T2D) remains elevated, affecting approximately 11.3% of the U.S. population and 9.3% of individuals globally, including minority groups^1,2^. Present clinical treatment guidelines target achievement of glycated hemoglobin A1c (HbA1c) levels below 7% for most adults^3,4,5^. Metformin (MF) remains the first-line of therapy, however, newer second-line treatments are often needed when metformin alone fails to achieve glycemic control^6–9^. Factors, particularly social determinants of health (SDOH), which are the conditions in the environments where people are born, live, learn, play, worship, and age are known to impact a range of health, functioning, and quality of life outcomes^9,10^. Specific SDOH^10^ such as neighborhood disadvantage and socioeconomic status, along with other adverse factors, have been previously studied and shown to significantly impact diabetes management^11–14^, medication adherence^15^, and behaviors such as physical activity and dietary patterns^16–23^. Real-World characterization studies show wide-spread heterogeneity in the choice of second-line treatments^24–26^. However, the knowledge about the possible influence of patient-level SDOH on the allocation of second-line treatments among patients with T2D remains limited

We sought to systematically analyze the association between SDOH, specifically socioeconomic status (SES) as estimated by mapping of home location to the Area Deprivation Index (ADI)^27^ and the allocation of second-line treatments among patients with T2D after MF monotherapy. The study leveraged clinical data from electronic health records (EHRs) of over 9 million patients across five academic medical centers within the University of California Health (UC Health) system.

## Methods

### Data

Data for this study was extracted from the University of California (UC) Health System, which includes 20 health professional schools (6 medical schools), 5 academic health centers (UC San Francisco, UC Los Angeles, UC Davis, UC Irvine, and UC San Diego), and 12 hospitals. It is the eighth largest non-profit health system in the United States by revenue and has built a secure central data warehouse (UCHDW) for operational improvement, promotion of quality patient care, and enabling the next generation of clinical research^28^. The repository currently holds data securely on over 9 million patients seen since 2012. Every quarter, EHR data is extracted from each site and transformed into vendor neutral Observational Medical Outcomes Partnership (OMOP) common data model used widely across the United States and the world29. De-identification of the data has been completed to enable clinical research projects, under guidance from UC campus institutional review boards, privacy, and compliance. Research use of UCHDW is considered non-human subject’s research.

### Study Population

UCHDW data was extracted from patients diagnosed with T2D and had a documented prescription of second-line treatment in addition to existing MF therapy. Patients with T2D were identified using ICD-10 diagnosis code E11.*, HbA1c lab measurements **(eMethods Supp. 2),** and T2D related drug prescriptions for four second-line treatment classes (GLP-1RA, SGLT2i, DPP4i, and SU) **(eMethods Supp. 1),** from EHRs spanning from year 2012 up to November 2023. To select the cohort of patients who received a second-line SU prescription, for example, we excluded individuals who had a previous order of another second-line treatment prior to the prescription for an SU and those with documented type 1 diabetes or an atypical form of diabetes such as gestational diabetes. We also excluded patients who did not have an order for MF before receiving an SU, and those who lacked T2D diagnosis code prior to receiving the SU. We also required patients to have documented HbA1c measurements within (+7/-7d) of SU treatment initiation (index date), where the index date was the first prescription of SU (**Figure-1**). Finally, we excluded all patients whose HbA1c was < 6.5% and were under 17 years of age at the time of SU treatment initiation and those who did not have an ADI value reported at the index date. Similar exclusion and inclusion criteria were applied to select individuals with other second-line T2D medications. This study followed the Strengthening the Reporting of Observational Studies in Epidemiology (STROBE) reporting guidelines to ensure the quality of our observational study^30^.

**Figure 1:**
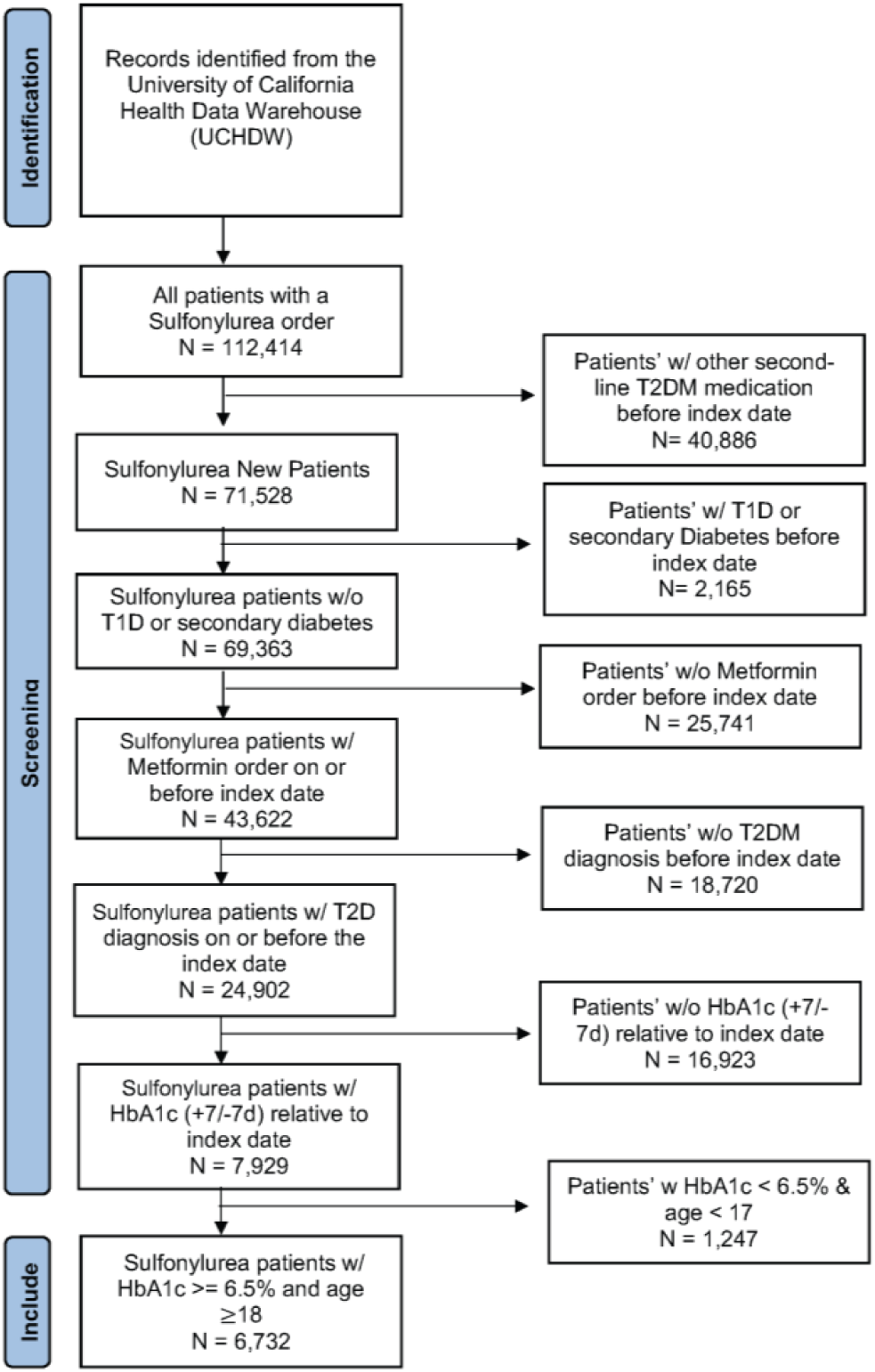
Second-Line Treatment Cohort Design. Flow chart illustrating cohort selection for patients being started on second-line sulfonylurea (SU) across five academic health systems in University of California Health. A similar cohort selection process was repeated for each of the other 3 categories of second-line treatments.

### Area Deprivation Index (ADI)

We utilized the ADI as a metric to estimate patient-level SDOH^27^. ADI serves as a valuable resource for identifying and assessing healthcare needs for clinical treatments among patients by providing a general estimate of SDOH based on home location census tract characteristics. This index integrates SDOH data from 17 different census tract variables sourced from the U.S. Census, with factors related to poverty, housing, employment, and education. ADI as a metric has been used in previous observational studies^31–35^ to understand its SDOH role on health outcomes in other disease states. For this study, census tract-based SDOH data was calculated to create the ADI, which ranks areas by percentiles, distinguishing the most deprived areas with higher percentile scores from the least deprived areas with lower percentile scores^27,36,37^.

### Study Variables

To describe patient baseline characteristics, we used ADI, HbA1c, Age (in years), patient reported Sex (Male, Female), presence of primary insurance coverage, patient-reported Race, and Ethnicity. T2D severity was inferred as the average of HbA1c measurements taken within the (+7/-7d) window relative to the index date. Age (in years) was calculated relative to the index date from date of birth. Patients with primary insurance healthcare coverage were utilized as Medicare, Medicaid, or private insurance types. Patient reported race was categorized as White, Asian, Black or African American category, Native Hawaiian or Pacific Islander, American Indian or Alaska Native category, Multi-Race, Other Race, or Unknown. Patient reported ethnicity was categorized as Hispanic or Latino, Not Hispanic or Latino or Unknown.

### Statistical Analysis

Descriptive statistics were calculated first for each covariate and then for the overall patient population. Multinomial regression was used to estimate the association of ADI with second-line treatment allocation, adjusting for other covariates considered in this study^38,39^. For the multinomial regression, the four second-line treatment classes (GLP-1RA, SGLT2i, DPP4i, and SU) were considered as outcomes, with SU serving as the reference class. We adjusted for HbA1c, age (in years), sex (Male, Female), primary insurance coverage, race, and ethnicity by using a multinomial regression model ^40,41^. Association and its strength was reported as odds ratio with 95% confidence Intervals (CI) including p-values indicating the statistical significance of how likely a patient was to receive a second-line treatment compared with SU when adjusted for study covariates. An Odds Ratio (OR) was considered significant if it’s 95% CI did not span 1, and p-value < 0.05. An OR = 1 indicates no difference between SU and other second-line treatments, OR > 1 indicates an increased likelihood of allocating other second-line treatments compared with SU, and OR < 1 indicates decreased likelihood of allocating other second-line treatments compared with SU. All calculations were performed using R statistical software version 3.6.3 (R Project for Statistical Computing).

### SDOH Identification through Large Language Models

To identify individual-level SDOH factors, we extracted T2D patients from the UCSF Deidentified clinical database using the inclusion and exclusion criteria outlined in the study population section of the methods. We excluded patients who did not have clinical notes associated with their diagnoses. Further, we excluded patients whose clinical notes did not contain Social History or SDOH information, resulting in a final cohort of 105 patients with 270 clinical notes **(Supplement-3)**. To identify individual-level SDOH factors, we utilized the GPT-4 inference model with a zero-shot learning approach. GPT-4 was prompted to analyze the Social History and SDOH sections of the clinical notes written by physicians. The model’s accuracy was first tested and validated using an annotated synthetic dataset and further evaluated with an annotated MIMIC-III dataset curated by Guevara et al^42^. Outputs from these sample datasets were compared against the ground truth to check for accuracy in identifying SDOH factors **(Supplement-3)**. GPT-4 was then prompted with the task to identify SDOH factors within UCSF clinical notes, categorized into the following domains as outlined in the annotation guidelines by Guevara et al.: Transportation (Distance, Resource, Other), Housing (Poor, Undomiciled, Other), Relationship (Married, Partnered, Divorced, Widowed, Single), Parent, Support (Minus, Plus), Employment (Employed, Underemployed, Unemployed, Disability, Retired, Student)^42^. Adverse SDOH factors were consolidated into broader categories: Adverse Transportation, Adverse Housing, Adverse Relationship, Patients with Children, Adverse Support, and Adverse Employment. The identified SDOH factors were visualized and plotted.

## Results

Our study included 15,090 patients (mean age 59.3 years, SD 13.4 years). **Table 1** presents a summary of treatment allocation and demographic information. Of these patients, 6,732 were prescribed SU, 2,704 were prescribed DPP4i, 2,918 were prescribed GLP-1RA, and 2,736 were prescribed SGLT2i after final inclusion and exclusion criteria. 56.7% of the patients identified as male, 43.3% female, and 46.6% identified as White. Primary Insurance coverage was distributed among patients as private (43.1%), medicaid (30.1%), medicare (25.0%), and veterans affairs (1.8%). The mean [SD] HbA1c values, reflecting T2D disease severity across second-line treatments, were: DPP4i (8.59% [1.85]), GLP-1RA (8.83% [2.06]), SGLT2i (8.62% [1.88]), and SU (9.19% [2.02]). Overall, the ADI for each second-line treatment allocation was skewed toward the least economically disadvantaged neighborhoods (ADI < 5), totaling 52.4% of patients in the study cohort. Older patients (OR = 1.02 [95% CI, 1.01-1.02]), as well those self-identifying as Asian (OR = 1.19, [95% CI, 1.05-1.35]), American Indian or Alaskan Native (OR = 1.33, [95% CI, 0.74-2.41]) were more likely to be prescribed a DPP4i as compared to individuals who identify as White (**Table 2**) adjusting for ADI. Females (OR = 1.44, [95% CI, 1.31-1.58]) were more likely to be prescribed GLP-1RA compared to Males. Patients who identified as Unknown Race (OR = 1.81, [95% CI, 1.49-2.20]) were more likely to be prescribed SGLT2i when compared to Whites. While patients identifying as “Other” race (OR = 1.21, [95% CI, 1.06-1.38]) were more likely to be prescribed DPP4i when compared to White patients. Hispanic patients were less likely to be prescribed DPP4i (OR = 0.73, [95% CI, 0.64-0.83]), GLP-1RA (OR = 0.78, [95% CI, 0.69-0.88]), and SGLT2i (OR = 0.90, [95% CI, 0.79-1.02]) when compared to non-Hispanic patients.

**Table 1:**
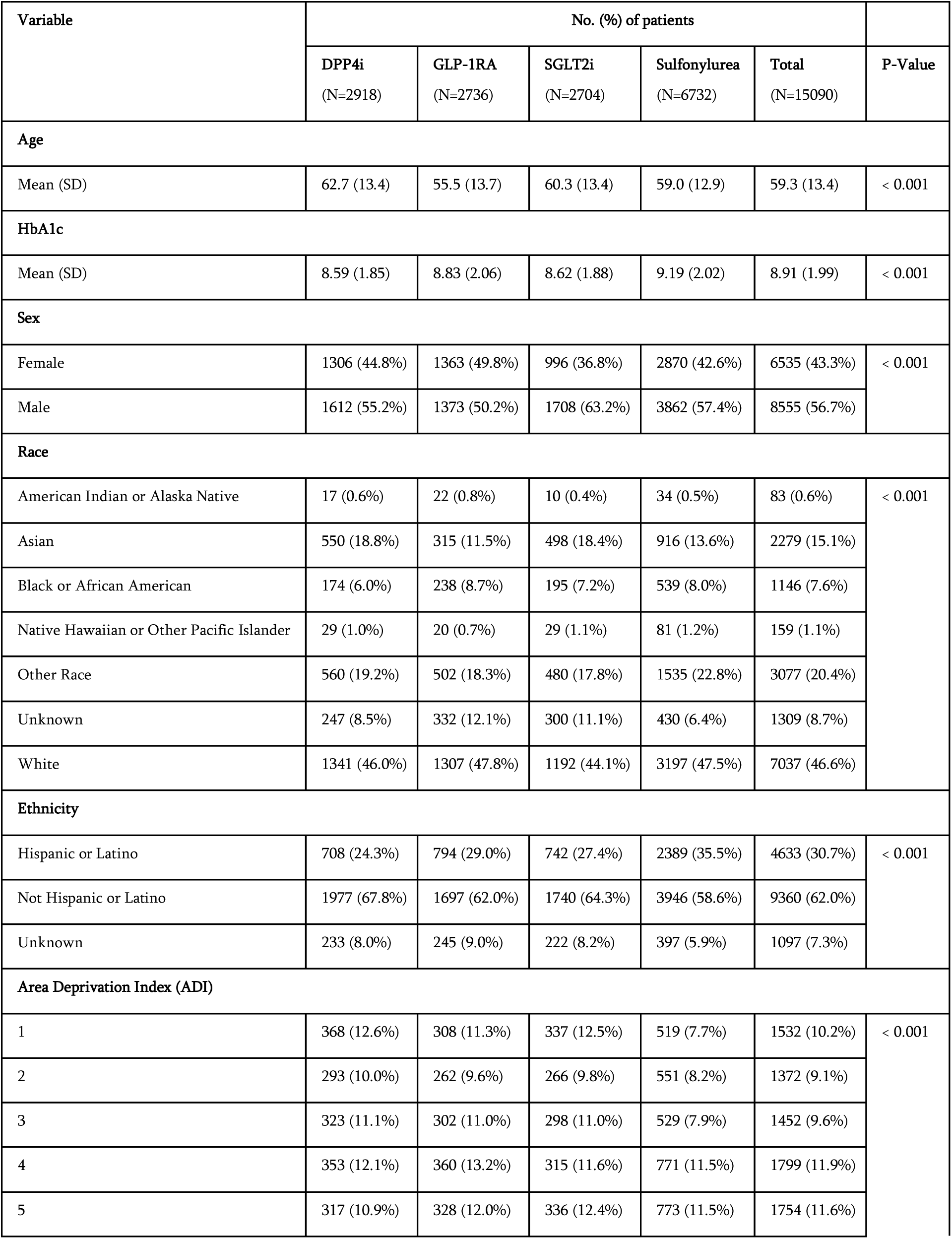
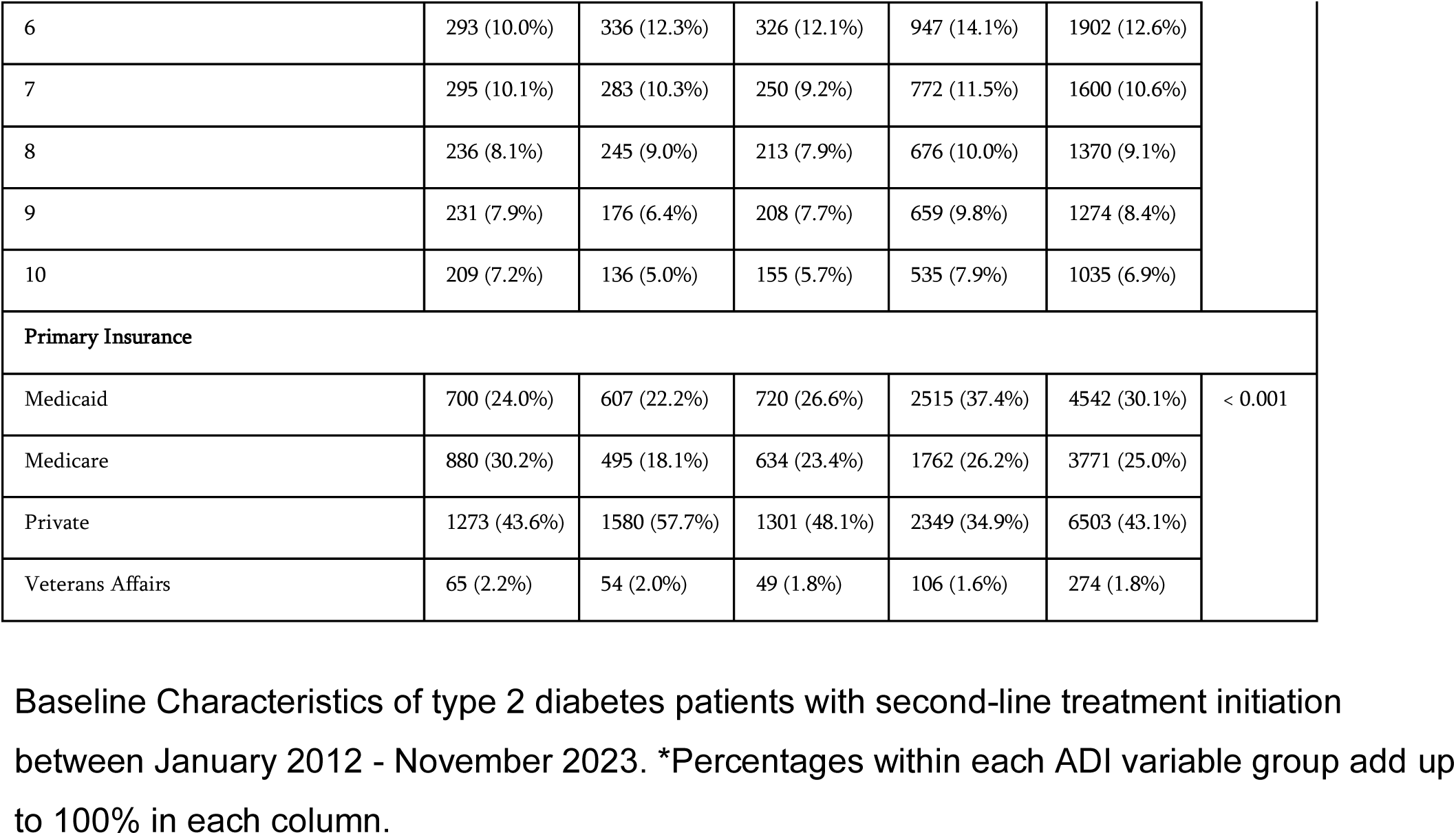
Baseline Characteristics of Type 2 Diabetes Patients with Second-Line Treatments.

**Table 2:** Results of the Multinomial Regression Model.

| Variable | DPP4i |  | GLP-1RA |  | SGLT2i |  |
| --- | --- | --- | --- | --- | --- | --- |
|  | OR (95% CI) | P value | OR (95% CI) | P value | OR (95% CI) | P value |
| **Area Deprivation Index (ADI)** | 0.96 (0.95-0.98) | < 0.001 | 0.94 (0.92-0.96) | < 0.001 | 0.95 (0.93-0.97) | < 0.001 |
| **HbA1c** | 0.90 (0.88-0.92) | < 0.001 | 0.90 (0.87-0.92) | < 0.001 | 0.88 (0.85-0.90) | < 0.001 |
| **Age** | 1.02 (1.01-1.02) | < 0.001 | 0.98 (0.97-0.98) | < 0.001 | 1.01 (1.00-1.01) | 0.001 |
| **Sex** |  |  |  |  |  |  |
| Female | 1.10 (1.00-1.20) | 0.043 | 1.44 (1.31-1.58) | < 0.001 | 0.79 (0.72-0.86) | < 0.001 |
| **Race** |  |  |  |  |  |  |
| American Indian or Alaska Native | 1.33 (0.74-2.41) | 0.345 | 1.52 (0.87-2.64) | 0.138 | 0.85 (0.42-1.74) | 0.662 |
| Asian | 1.19 (1.05-1.35) | 0.008 | 0.62 (0.53-0.72) | < 0.001 | 1.25 (1.09-1.43) | 0.001 |
| Black or African American | 0.81 (0.67-0.98) | 0.031 | 1.03 (0.86-1.22) | 0.759 | 1.06 (0.88-1.27) | 0.533 |
| Native Hawaiian or Other Pacific Islander | 0.86 (0.56-1.33) | 0.507 | 0.53 (0.32-0.87) | 0.012 | 1.01 (0.65-1.56) | 0.969 |
| Other Race | 1.21 (1.06-1.38) | 0.004 | 1.04 (0.91-1.19) | 0.552 | 1.04 (0.90-1.19) | 0.604 |
| Unknown | 1.35 (1.10-1.65) | 0.004 | 1.54 (1.27-1.86) | < 0.001 | 1.81 (1.49-2.20) | < 0.001 |
| **Ethnicity** |  |  |  |  |  |  |
| Hispanic or Latino | 0.73 (0.64-0.83) | < 0.001 | 0.78 (0.69-0.88) | < 0.001 | 0.90 (0.79-1.02) | 0.104 |
| Unknown | 1.02 (0.83-1.25) | 0.859 | 0.92 (0.75-1.14) | 0.459 | 0.90 (0.73-1.12) | 0.346 |
| **Primary Insurance** |  |  |  |  |  |  |
| Medicare | 1.19 (1.05-1.36) | 0.007 | 1.30 (1.12-1.51) | < 0.001 | 0.93 (0.81-1.07) | 0.297 |
| Private Health Insurance | 1.67 (1.49-1.87) | < 0.001 | 2.39 (2.13-2.69) | < 0.001 | 1.56 (1.40-1.75) | < 0.001 |
| Veterans Affairs | 1.62 (1.17-2.24) | 0.004 | 2.26 (1.60-3.21) | < 0.001 | 1.29 (0.91-1.84) | 0.158 |

Odds Ratio table of patients receiving second-line treatment across patient characteristics with SU as the reference. An OR = 1 indicates no difference between SU and other drugs, OR > 1 indicates increased likelihood of other treatments being used as a second line therapeutic compared with SU and OR < 1 indicates decreased likelihood of other treatments compared with SU.

Our results also highlighted that HbA1c was a significant determinant of second-line treatment allocation, specifically preferring an order of SU after metformin initiation, aligning with known treatment guidelines^6^. With every unit increase in HbA1c percentage indicative of increased disease severity, patients were more likely to be prescribed an SU versus any of the other three second-line treatments DPP4i (OR = 0.90, [95% CI, 0.88-0.92]), GLP-1RA (OR = 0.90, [95% CI, 0.87-0.92]), SGLT2i (OR = 0.88, [95% CI, 0.85-0.90]). Additionally, our results highlighted that primary insurance coverage was significant in determining second-line treatment allocation. Patients with Medicaid as their primary insurance coverage were less likely to receive to GLP-1RA when compared to patients with private (OR = 2.39, [95% CI, 2.12-2.69]), medicare (OR = 1.30, [95% CI, 1.12-1.51]), or veterans affairs insurance (OR = 2.26, [95% CI, 1.60-3.21]).

Interestingly, we found ADI to be a significant independent variable underlying allocation of four second-line treatments. As ADI increased from a relatively low value reflecting a least economic disadvantage to progressively higher values reflecting low socioeconomic neighborhoods, patients were more likely to be prescribed an SU versus the other three second-line treatments DPP4i (OR = 0.96, [95% CI, 0.95-0.98]), GLP-1RA (OR = 0.94, [95% CI, 0.92-0.96]), SGLT2i (OR = 0.95, [95% CI, 0.93-0.97]). As illustrated with randomly selected patients that followed the similar inclusion and exclusion criteria in our study but were not part of the model building (**Figure-2**), a Hispanic female patient aged 46 years with an HbA1c of 11.8% and Private insurance as their primary insurance living in a least disadvantaged neighborhood (ADI 1) is most likely to be prescribed a GLP-1RA as a second-line medication in addition to MF (38.5% likelihood). Whereas a non-Hispanic White male patient aged 49 years with an HbA1c of 11.9% and Medicaid as their primary insurance who lives in a economically disadvantaged neighborhood (ADI 10) is most likely to be prescribed SU as a second-line treatment (67.2% likelihood). In another example, a Hispanic female patient aged 53 years with a 9.2% HbA1c and Medicaid insurance living in ADI 10 is most likely to be prescribed SU as a second-line treatment (61.0% likelihood).

**Figure 2:**
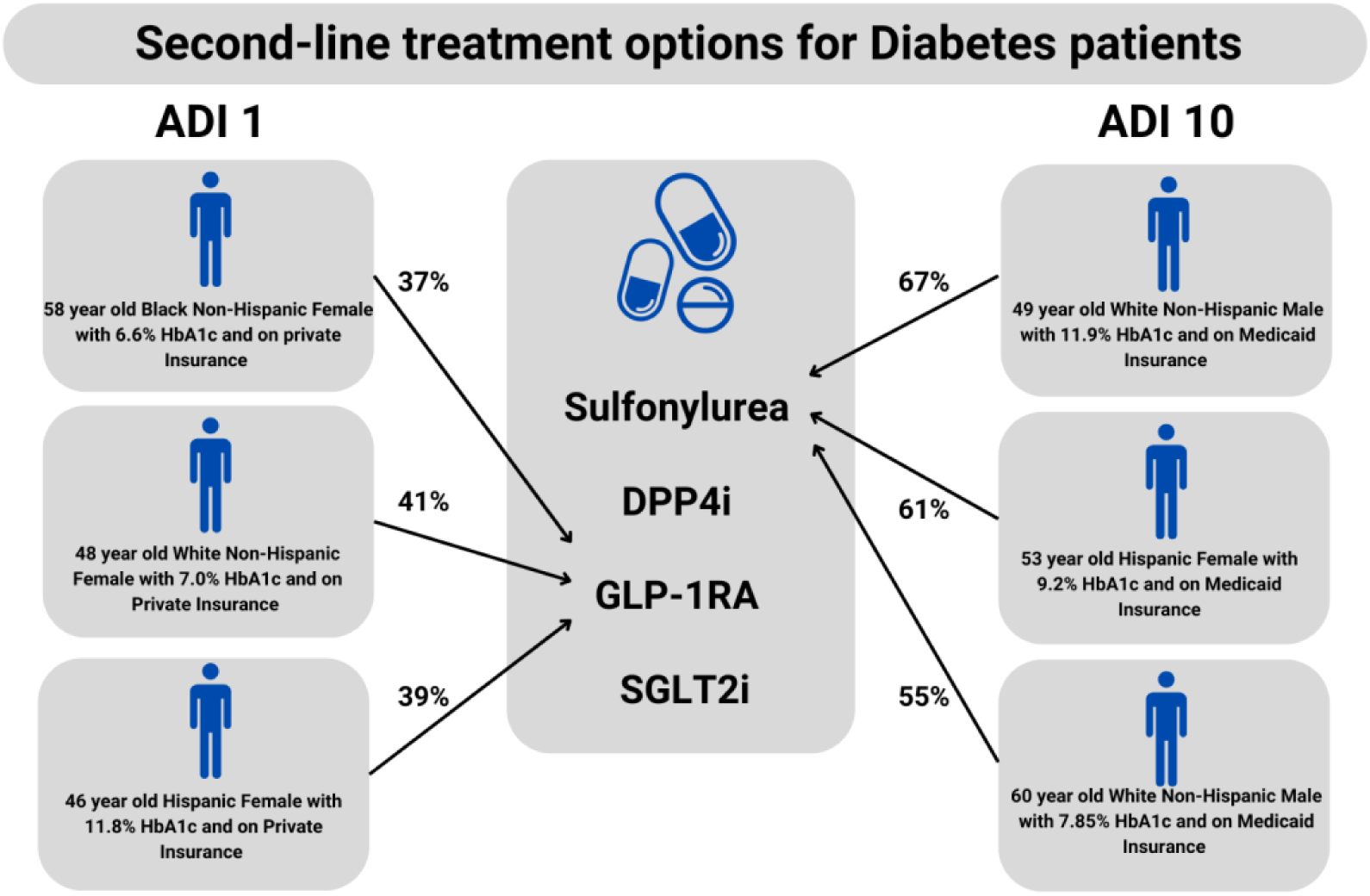
Second-line treatment options for Type 2 diabetes patients. Patients randomly selected from our cohort with varying characteristics (age, race, ethnicity, HbA1c, and insurance) are shown here. When each of these patients lives in an economically advantaged neighborhood (ADI 1, on the left) the model determines that they are most likely to receive a GLP-1RA as a second line treatment (percent indicates the model likelihood). When those same patients live in an economically disadvantaged neighborhood (ADI 10, on the right), they are most likely to receive a sulfonylurea.

Our results from prompt engineering using GPT-4 LLMs showed the identification of individual-level SDOH factors for 105 patients whose clinical notes contained social history. Among UCSF patients with clinical notes, 54 were prescribed SU, 21 were prescribed DPP4i, 15 were prescribed GLP-1RA, and 15 were prescribed SGLT2i. We identified the following present SDOH factors among these patients: adverse housing (n=8), adverse transportation (n=1), adverse relationships (n=22), adverse employment (n=12), adverse support (n=1), and patients with children (n=25) **(Figure-3)**.

**Figure 3:**
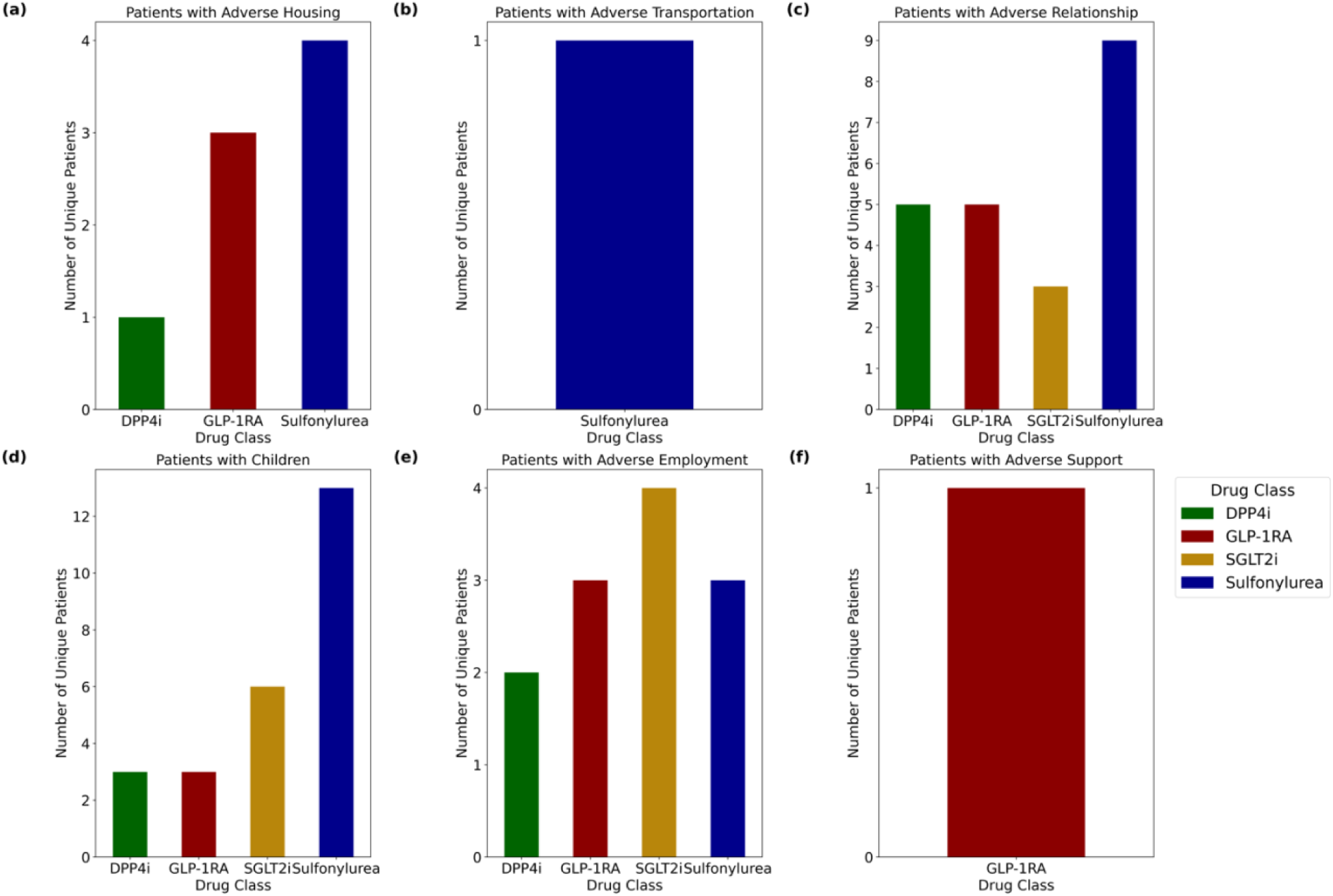
GPT-4 extraction and identification of Individual-Level SDOH Factors. GPT-4 was provided with 522 clinical notes with Social History and SDOH sections and was prompted to extract and identify individual-level SDOH factors from patients with a second-line T2D treatment. Identified Individual-Level SDOH Factors for UCSF patients with 3a. Adverse Housing 3b. Transportation, 3c. Relationship, 3d. patients with children, 3e. Employment, and 3f. Support stratified by their Drug Class.

## Discussion

In this retrospective cohort study, we evaluated the association of social determinants of health such as ADI on the allocation of second-line treatments among patients with T2D. We found that as socioeconomic disadvantage metrics ranged from relatively affluent to progressively socioeconomically disadvantaged neighborhoods, patients were more likely to receive an SU, a less clinically effective, but more cost-effective treatment^3,6,8^. Conversely, patients who lived in relatively least socioeconomic disadvantaged neighborhoods were more likely to receive newer, more expensive, and potentially more effective second-line treatments such as SGLT2i, DPP4i, and GLP-1RA. We also found that patients with Medicaid as their primary insurance, often associated with low-cost healthcare coverage, were less likely to receive these advanced second-line therapies. This disparity underlying treatment allocation is important, given that SGLT2i and GLP-1RAs have clearly shown benefits in limiting development of serious comorbid conditions including CKD, cardiovascular events, heart failure, and obesity^3,6^. Our findings thus highlight a gap in the fair allocation of effective second-line treatments among patients with T2D.

Beyond underlying socioeconomic disparities in second-line treatment allocation, we also found a reliance favoring SU when the urgency for glucose lowering was highest. Indeed, in each comparative analysis of specific agents as second-line treatments for T2D, we found that the worse a patient’s glycemic control, as measured by HbA1c, the more likely that patient was to receive an SU. This reliance is in line with the traditional prescribing patterns and older ADA guidelines that emphasize SU treatment as the first adjunct to MF for T2D. It also highlights the notion that older more established medication classes might be favored when A1c lowering is the top priority, regardless of SES. This fits with classic literature and ongoing supportive data indicating that optimal HbA1c lowering remains the most important factor in limiting the development of T2D-associated complications^6^. This finding is important because it highlights that patients with a high ADI are more likely to have a more severe A1c at second-line treatment initiation and are more likely to receive SU.

Our analysis also revealed disparities in second-line treatment allocation based on patient sex and age. First, we found that males were more likely to receive a DPP4i, or a GLP-1RA over SU compared to females, potentially consistent with the fact that adult males have higher T2D rates than females^43^. Advancing age was associated with a higher likelihood of receiving an SU over other second-line treatment choices. This finding may reflect a preference for SU among aged patients to minimize risk of hypoglycemia, cognitive impairment, and autonomic symptoms during management of diabetes^6^.

Our study showed that patients prescribed SU as a second-line treatment had a higher prevalence of documented adverse SDOH factors compared to those prescribed GLP-1RA, SGLT2i, and DPP4i. More specifically, these patients faced challenges such as adverse housing, relationships, transportation issues, with many also having caregiving responsibilities for children. This suggests they may have been prescribed a cost-effective treatment rather than a newer second-line option due to non-medical factors. Interestingly, patients prescribed SGLT2i had a higher prevalence of documented adverse employment factors than those prescribed other treatments, indicating that some patients facing employment challenges are still being treated with newer drugs. However, the relatively low number of individual-level SDOH factors captured in our data provides only a partial view of the adverse contributors influencing a patient’s second-line treatment prescription.

There are several limitations in the study. The health disparities associated with SES in the UCHDW, as highlighted in our study, were estimated using the ADI at the neighborhood level, which only provides an approximate measure of a patient’s SES. Additionally, the inability to capture a significant amount of individual-level SES data and other SDOH parameters due to the absence of such data elements, limited a comprehensive evaluation of the patient population. We acknowledge that our understanding of SES at the individual level remains incomplete and could potentially be improved in future studies. The sample size of our data was also constrained, primarily due to the stringent cohort selection criteria employed in this study.

Furthermore, our study does not account for the approval dates of newer drugs such as SGLT2i, DPP4i, and GLP-1RA which were introduced later during the study period. Lastly, the distribution of patients in this study was slightly skewed towards least economically disadvantaged neighborhoods due to the high number of patients living in higher socioeconomic areas that are treated within the UC-Health System.

Our findings serve as a preliminary step toward a deeper exploration of how individual SES and SDOH can contribute to health disparities when data is collected in large quantities. We believe that differences in disease severity at second-line treatment allocation may stem from factors such as limited access to healthcare, financial constraints, low health literacy, and geographic location of a patient. Moreover, our findings suggest that disparities in treatment allocation may be influenced by treatment cost and unequal access to healthcare providers who prescribe newer, more effective medications (e.g., GLP-1RA). To further understand the disparities highlighted in our study, a comprehensive examination of high-quality individual-level SDOH factors is needed to effectively address the underlying issues pertaining to treatment selection and disease severity disparities.

In conclusion, our analysis highlights the association of SDOH with treatment choices, significantly impacting disease state and health outcomes. Our study shows growing associations of SDOH on clinical treatments, with and without adjusting for other covariates. Furthermore, our study shows that SDOH can be utilized to understand differences in care among patients from distinct population groups based on socioeconomic status. The recognition of SDOH disparities within the UC-Health system for T2D extends to healthcare systems across the United States. Understanding the role of SDOH, alongside considerations of disease severity, age, and sex, is crucial in addressing disparities in care among diverse population groups. A comprehensive understanding of treatment disparities will lead to the fair allocation of medical resources to population groups in which disparities exist. Most importantly utilizing SDOH to understand differences in care for multiple disease could drive institutions to improve their delivery of care to disparate population groups. Therefore, we must encourage the generation of high-quality SDOH data at both the neighborhood and individual level to increase the viability of clinical-related questions across disease states.

## Data Availability

All data produced in the present study are not available to the public.

## Author Contributions/Acknowledgements

Conceptualization, J.D., R.V., K.R and A.J.B.; writing—original draft preparation, J.D.; writing— review and editing, J.D., R.V., S.K.K, and A.J.B, Area Deprivation Index data, A.P. All authors have read and agreed to the published version of the manuscript.

